# Genotype-Phenotype Correlations Reveal Positive Inheritance and Phenotype Associations for *PROM1*-Associated Inherited Retinal Degenerations

**DOI:** 10.64898/2026.09.03.26361572

**Authors:** Mahnoor Shoukat, Kimberly M. Papp, Ehsan Misaghi, Danan J. D. Kalra, Ian M. MacDonald, Matthew D. Benson, Brittany J. Carr

## Abstract

**Objective:** We analyzed 190 PROM1 variants to determine whether we could identify genotype-phenotype correlations with predictive value for patient outcomes. We present a case-series of 7 patients with rare, under-reported, or unique forms of *PROM1*-associated retinal dystrophies.

**Design:** We performed a retrospective database study by searching for human *PROM1* variants reported to be pathogenic, likely pathogenic, or disease-causing in two online databases: ClinVar and Human Gene Mutation Database. Contingency tables were constructed variable pairs – inheritance pattern vs. variant type, inheritance pattern vs. reported phenotype, inheritance pattern vs. protein domain, phenotype vs. variant type, phenotype vs. protein domain, and variant type vs. protein domain – and were then analyzed using a Monte Carlo chi-square test of independence with adjusted standardized residuals. Cells with absolute standardized residuals ≤ -1.96 and ≥ 1.96 were interpreted as under- or over-represented relative to expectation (p < 0.05).

**Participants:** Seven participants with *PROM1*-associated retinal degeneration were contacted from our ocular genetics clinical practice; all contacted patients or legal guardians consented to be included in this case-series.

**Main Outcome(s) and Measure(s):** The primary study outcome was to determine whether there were any meaningful genotype-phenotype relationships for disease-causing *PROM1* variants by chi-square correlation analysis. Patient case-series were included to enrich statistical findings.

**Results:** We found a meaningful increase in observed counts of autosomal dominant missense variants associated with macular dystrophy. Of 7 patients, 3 were women (43%) and ages ranged from 4-58 years. Four of the seven patients had dominantly inherited missense variants (c.1117C>T, c.1557C>A, c.2110C>T, c.1655T>C). Recessive patient variants were all predicted to generate a truncated protein, including two patients with the same variant (c.1423_1424del), and a patient with a c.1354dup variant and compound heterozygous *ABCA4* variants of unknown significance (c.2382+95A, c.-79C>T). All patient phenotypes were in agreement with predicted phenotypic outcomes from our statistical analysis.

**Conclusions:** We provided statistical outcomes and patient case-series data that support predictive value for dominant and recessive forms of *PROM1*-associated inherited retinal degeneration. These data could inform expected outcomes for patients with *PROM1-*associated blindness, lessoning anxiety about unknown outcomes, and provide a framework to inform best-practices for patient follow-up.

---

Inherited retinal degeneration describes a genetically diverse range of conditions that result in progressive photoreceptor cell death and blindness. Inherited retinal degeneration affects 1:1500-1:30,000 people in North America, and although they are individually considered rare diseases, combined, they are a significant source of blindness^1,2^. Prominin-1 (*PROM1*; OMIM 604365) is a associated with inherited blindness and the resultant phenotypes have remarkable phenotypic heterogeneity, including retinitis pigmentosa, rod-cone dystrophy, cone-rod dystrophy, Stargardt-like disease, and Leber congenital amaurosis (LCA); it can also be inherited in an autosomal dominant or autosomal recessive fashion^3–7^.

PROM1, also known as AC133/CD133, was first identified in human progenitor and hematopoietic stem cells^8,9^. Later, it was found in fully differentiated tissues, including the kidneys, liver, brain, pancreas, placenta, mammary glands, and in the retina^8–10^. PROM1 is a transmembrane protein with an extracellular N-terminus, two large extracellular loops with nine N-linked glycosylation sites, two small intracellular loops, and an intracellular C-terminus (**Fig. 1A**). PROM1 is so named because it is concentrated in “prominent” membrane protrusions such as microvilli, microvilli-like protrusions of neuroepithelial cells, and membrane protrusions in non-epithelial cells such as filopodia, lamellipodia, micro spikes, and cilia^8^. In the retina, PROM1 is localized to lipid raft microdomains at the leading edges of outer segment membrane discs in the rod and cone photoreceptors (**Fig. 1B**)^5,11–13^. These lipid domains are rich in cholesterol and sphingolipids, and are known to be important for photoreceptor outer segment plasma membrane budding, growth, and organization^11,14–17^. In animal models with genetic modifications that affect PROM1 function, it appears to have an important role in maintaining photoreceptor outer segment morphology^5,11–13,18,19^. *Prom1^-/-^* mice, *prom1*-null frogs, and *prom1b^-/-^* zebrafish have severely dysmorphic rod and cone outer segments, and in fish and frogs, cones are more severely affected than rods^12,18–20^. *Prom1^-/-^* mice have thinning of the outer nuclear layer of the retina and displacement of outer segment proteins cone opsin, rhodopsin, photoreceptor cadherin (CDHR1) and rod outer segment membrane protein 1 (ROM1) into the photoreceptor inner segments, which is indicative of retinal degeneration^18,20–22^. A transgenic mouse model of autosomal dominant *Prom1*-associated disease (p.Arg373Cys) also have overgrown dysmorphic photoreceptor outer segments and mislocalization of CDHR1 in the inner segment^5^. Aging *prom1*-null frogs develop retinal deposits similar to subretinal drusenoid deposits (SDD), concomitant with retinal pigment epithelium (RPE) atrophy, loss of photoreceptor and bipolar cell synapses, and cone atrophy^12,23^. Thus, similar to humans, animal models demonstrate heterogeneity in *PROM1*-associated disease phenotypes.

**Figure 1.**
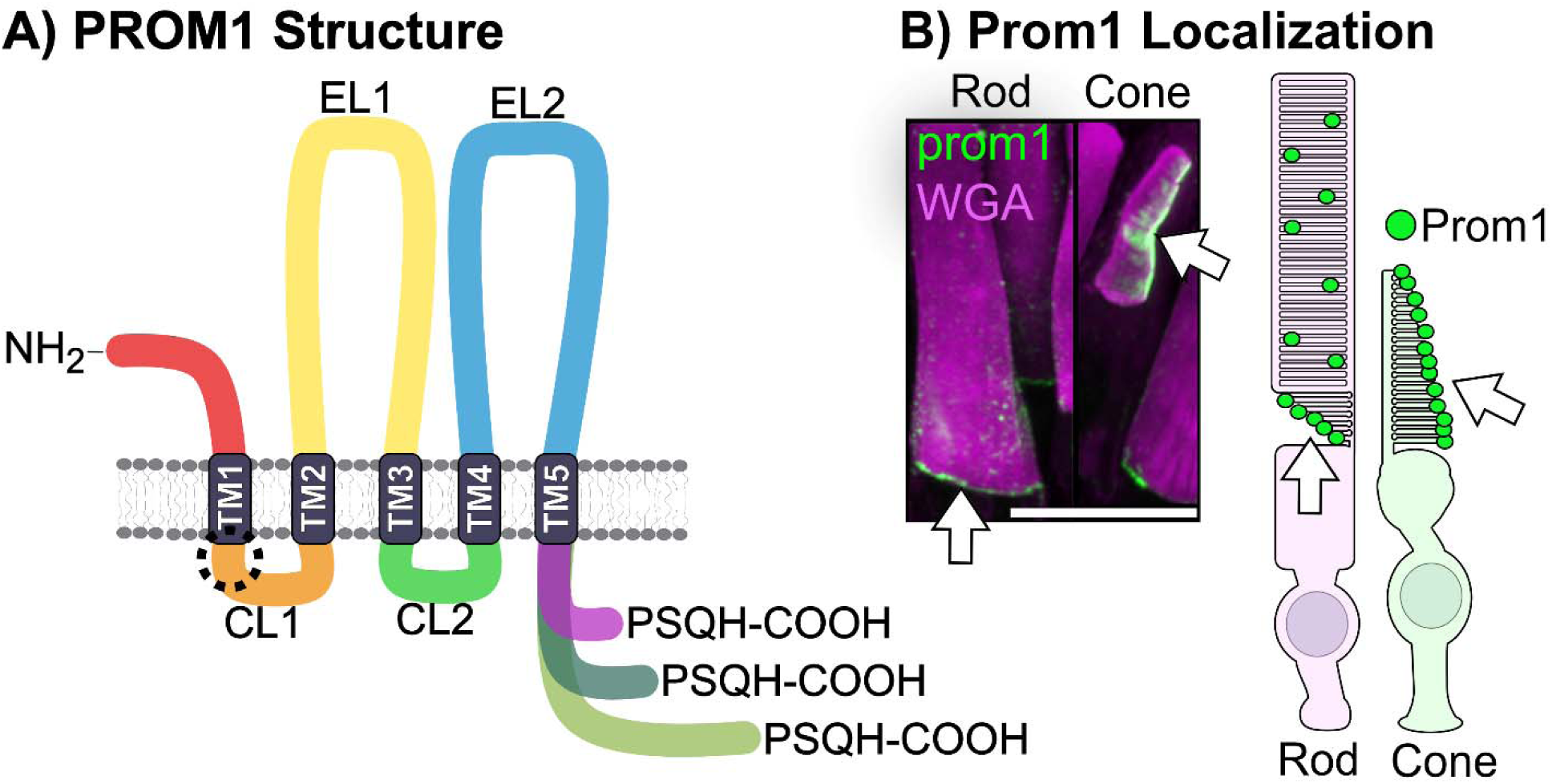
Retinal PROM1 protein structure and localization. A) PROM1 protein has an extracellular N-terminus (red), 5 transmembrane domains (grey), 2 cytoplasmic loops (CL1, orange and CL2, green), two large extracellular loops (EL1, yellow and EL2, blue), and a variably-spliced C-terminus (purple). B) Prom1 protein in the retina is localized to the open discs at the base of the rod outer segments and to the leading edges of the cone outer segments, opposite peripherin-2. Modified from Carr et al.

The heterogeneous nature of *PROM1*-associated blindness presents challenges in understanding the disease mechanism, identifying affected cellular pathways, identifying biomarkers, and controlling for potential biases in study design when performing genetic screens or investigating potential therapeutic pathways using clinical or foundational discovery science^24,25^. To address genotype/phenotype correlation in humans, we performed chi-square analysis on reported human *PROM1* pathogenic variants by their protein domain location, variant type, inheritance pattern, and phenotypic classification. We found strong, statistically-significant associations between dominant inheritance of missense variants and macular dystrophy, and a strong association between frameshift/nonsense variants and rod-cone dystrophy. We also report 7 *PROM1-*associated clinical case studies, including a rare pathogenic missense variant associated with cone dystrophy, a patient with a long (30+ year) natural history of *PROM1*-associated disease, two unrelated patients with the same variant, but differently assigned clinical phenotypes, a patient with a homozygous *PROM1* variant and compound heterozygous variants of unknown significance in *ABCA4*, a patient with a heterozygous *PROM1* missense variant with prominent drusen phenocopying age-related macular degeneration, and a patient with macular dystrophy caused by the heterozygous R373C variant for comparison.

## Methods

### Data Collection and Variant Categorization

We searched for reported variants in human PROM1 relevant to inherited blindness in two online databases: ClinVar (https://www.ncbi.nlm.nih.gov/clinvar/; accessed January 2025) and Human Gene Mutation Database (HGMD, https://www.hgmd.cf.ac.uk/ac/index.php; accessed January 2025). From ClinVar, we included variants categorized as pathogenic and likely pathogenic given their anticipated clinical relevance^26^. We used variants from HGMD categorized as disease-causing variants (DM). We excluded copy number variants such as large genomic deletions which involved multiple genes. Database-reported variant accuracy regarding phenotype and inheritance pattern was confirmed by cross-referencing original citations.

Each variant was categorized according to amino acid position, protein domain, variant type, inheritance pattern, and reported clinical phenotype(s). Variant type categorized by the effects that they were theorized to have on protein structure and function: 1) frameshift/nonsense 2) splice, 3) missense, and 4) in-frame deletion/duplication. Inheritance patterns were defined as: 1) recessive, 2) dominant, 3) conflicting, or 4) simplex/sporadic. The majority of inheritance patterns were unspecified (n = 98/190, 52%); these were included in the descriptive statistics, but not in the subsequent chi-square correlation analyses.

Clinical phenotypes were grouped into five broader categories to simplify categorization and statistics: 1) rod-cone dystrophy (RCD), 2) cone-rod dystrophy (CRD), 3) macular dystrophy (MD), 4) retinal dystrophy (RD), and 5) Leber Congenital Amaurosis (LCA). We defined RCD as including the additional term “retinitis pigmentosa”. We classified CRD as including the additional terms “cone dystrophy”. We classified macular dystrophy as including the additional terms “macular atrophy”, “macular degeneration”, and “Stargardt disease”. We classified retinal dystrophy as including the additional generalized terms “retinal disease”, “retinal degeneration”, and “retinal disorder”. No other search terms were included in the LCA category. Thirty-four of the 190 variants did not have a clinical phenotype classification. These were included in the descriptive statistics, but not in the subsequent chi-square correlation analyses.

### Statistical Analysis

To investigate associations between categorical variables, contingency tables were constructed for six variable pairs: inheritance pattern vs. variant type, inheritance pattern vs. reported phenotype, inheritance pattern vs. protein domain, phenotype vs. variant type, phenotype vs. protein domain, and variant type vs. protein domain. Analyses were restricted to cases with phenotypes classified as RCD, CRD, MD, RD, or LCA, and to variants with dominant or recessive inheritance. Contingency tables were analyzed using a Monte Carlo chi-square test of independence. A Monte Carlo test was used instead of a Pearson’s chi-square test because our data violated the core assumption of Pearson’s that at least 80% of cells must be ≥ 5, with a strict minimum of 1. A Monte Carlo chi-square overcomes these data limitations by generating thousands of random contingency tables with the same observed vs. expected margins, facilitating the construction of a precise null distribution via random sampling; we used the default number of simulations (n = 2000) for our analysis. To identify the specific pair combinations that had higher than expected observed counts, adjusted standardized residuals were also calculated. Cells with absolute standardized residuals ≤ -1.96 and ≥ 1.96 were interpreted as under- or over-represented relative to expectation (p < 0.05). Residuals for all variable pairs were visualized as heatmaps, with a diverging color scale where red indicates observed counts exceeding expected and blue indicates observed counts below expected. We then used a Cramér’s V test with bias correction to very chi-square results and measure the strength of any identified categorical associations. Variants, contingency tables, and standardized residuals are listed in **Supplementary Tables S1-S7**. All statistical analyses were conducted in R Studio (Version 4.6.0; 2026-04-24 ucrt). Heat maps were created using R Studio and the ggplot2 package (Version 4.0.3). All other figures were made using Affinity Designer (Version 2.6.5), and Affinity Photo (Version 2.6.5; Serif Ltd., West Bridgford, United Kingdom).

### Human Case Series

Human data were gathered under the approval of the University of Alberta Human Research Ethics Board (Pro00045377) and carried out in accordance with the Health Canada and Public Health Agency of Canada (PHAC) Research Ethics Board (REB), guided by the principles of the second edition of the Tri-Council Policy Statement: Ethical Conduct for Research Involving Humans (TCPS 2). This study was performed in accordance with the ethical standards as laid down in the 1964 Declaration of Helsinki and its later amendments.

Seven patients known to the University of Alberta Ocular Genetics service with disease-causing *PROM1* variants were contacted and enrolled in the case series. Each participant provided written informed consent to participate using large-print, technologically compatible forms. A retrospective chart review established clinical and investigative details including age of symptom onset, visual acuity, clinical phenotype, clinical diagnostic testing (including fundus photos, autofluorescence, and electroretinography results), and *PROM1* variant(s) identified through genetic testing. Patient 6 had a *PROM1* variant of unknown significance (VUS) (c.1655T>C), so the variant was not included in the database descriptives for pathogenic and likely pathogenic *PROM1* variants, but it was included in the protein domain and chi square analyses.

## Results

### Descriptive Statistics for Database Variants

We identified 190 unique disease-causing *PROM1* variants across ClinVar (92 pathogenic, 33 likely pathogenic) and HGMD (61 disease causing) databases (**Supplementary Table S1**). The majority of disease-causing *PROM1* variants were frameshift/nonsense variants (n = 103, 54%), followed by splice (n = 55, 29%), missense (n = 30, 16%), and then in-frame deletions/duplication variants (n = 2, 1%) (**Fig. 2A**). Of the five clinical phenotype categories analyzed, rod-cone dystrophy was the most commonly reported (n = 65, 34%), followed by cone-rod dystrophy (n = 34, 18%), retinal dystrophy (n = 34, 18%), not specified (n = 33, 17%), macular dystrophy (n = 21, 11%), and then finally LCA (n = 3, 2%) (**Fig. 2B**). The majority of variants had unspecified inheritance pattern (n = 97, 51%), but of the specified cases, autosomal recessive inheritance was the most common (n = 68, 36%), followed by dominant (n = 14, 7%), then conflicting (n = 6, 3%), and finally simplex/sporadic (n = 5, 3%)(**Fig. 2C**).

**Figure 2:**
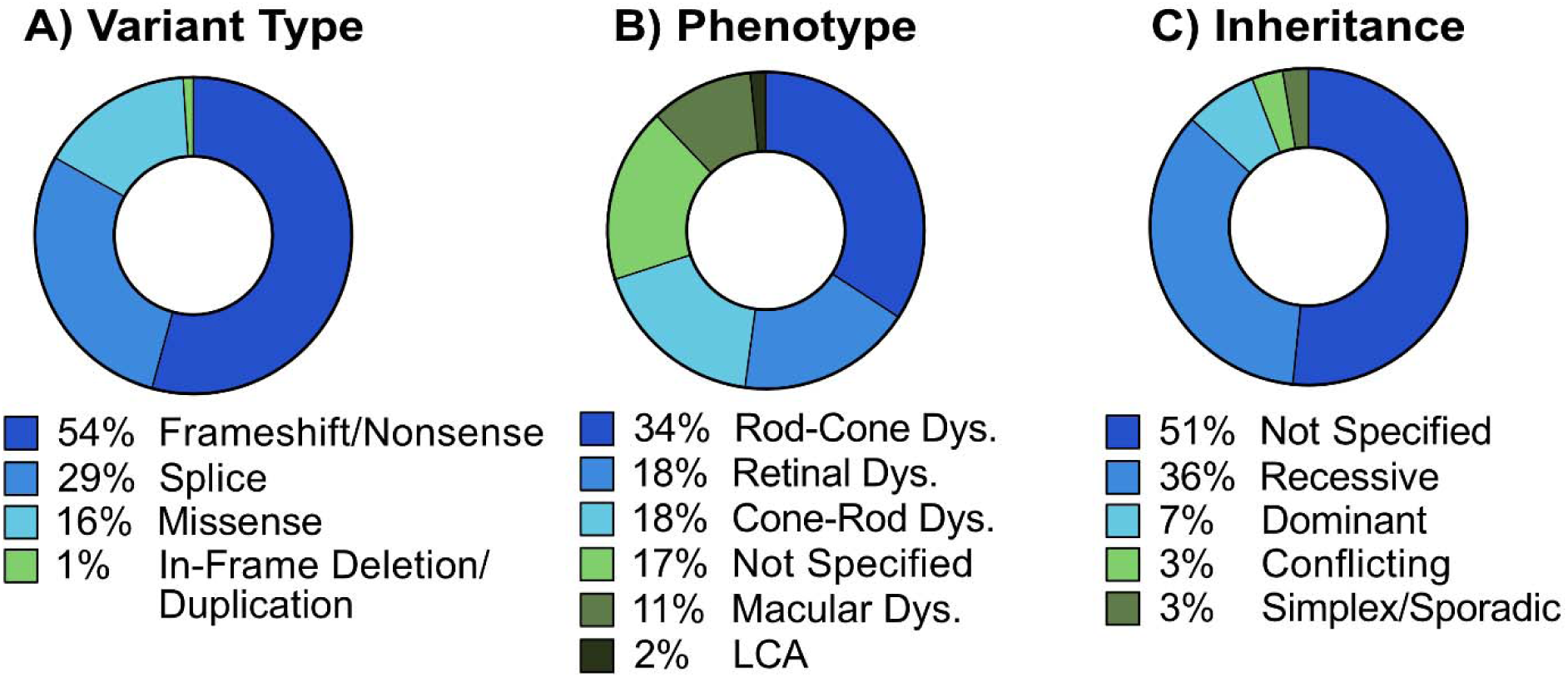
Descriptive statistics of the 190 reported PROM1 disease-associated variants collated from ClinVar and HGMD databases. (A) Variant types. (B) Clinical phenotype. (C) Inheritance patterns. *Abbreviations:* dys, dystrophy; LCA, Leber Congenital Amaurosis.

### Protein Domain Distribution of *PROM1* Variants

The three protein domains associated with the greatest disease-causing variant burden were extracellular loop 2 (n = 84, 44%), extracellular loop 1 (n = 42, 22%), and the N-terminus (n = 24, 13%; 79% of variants total). All other domains had less than 10% disease burden individually, and were as follows in protein domain order: transmembrane domain 1 (n = 3, 2%), cytoplasmic loop 1 (n = 6, 3%), transmembrane domain 2 (n = 3, 2%), transmembrane domain 3 (n = 8, 4%), cytoplasmic loop 2 (n = 10, 5%), transmembrane domain 4 (n =2, 1%), transmembrane domain 5 (n = 4), 2%, and the C-terminus (n = 5, 3%)(**Fig. 3**). When stratified by variant type, the N-terminus, extracellular loop 1, and extracellular loop 2 remained enriched for all outcomes. Regarding the patients reported in our case series, variant c.1345 dup (pTyr452Leufs*13) was found in transmembrane domain 3, variant c.1423_1424del (p. Val475Leufs*42) was found in cytoplasmic loop 2, and variants c.2110C>T (p.Arg704Cys) and c.1557C>A (p.Tyr519*) were found in EL2 (**Fig. 3**).

**Figure 3.**
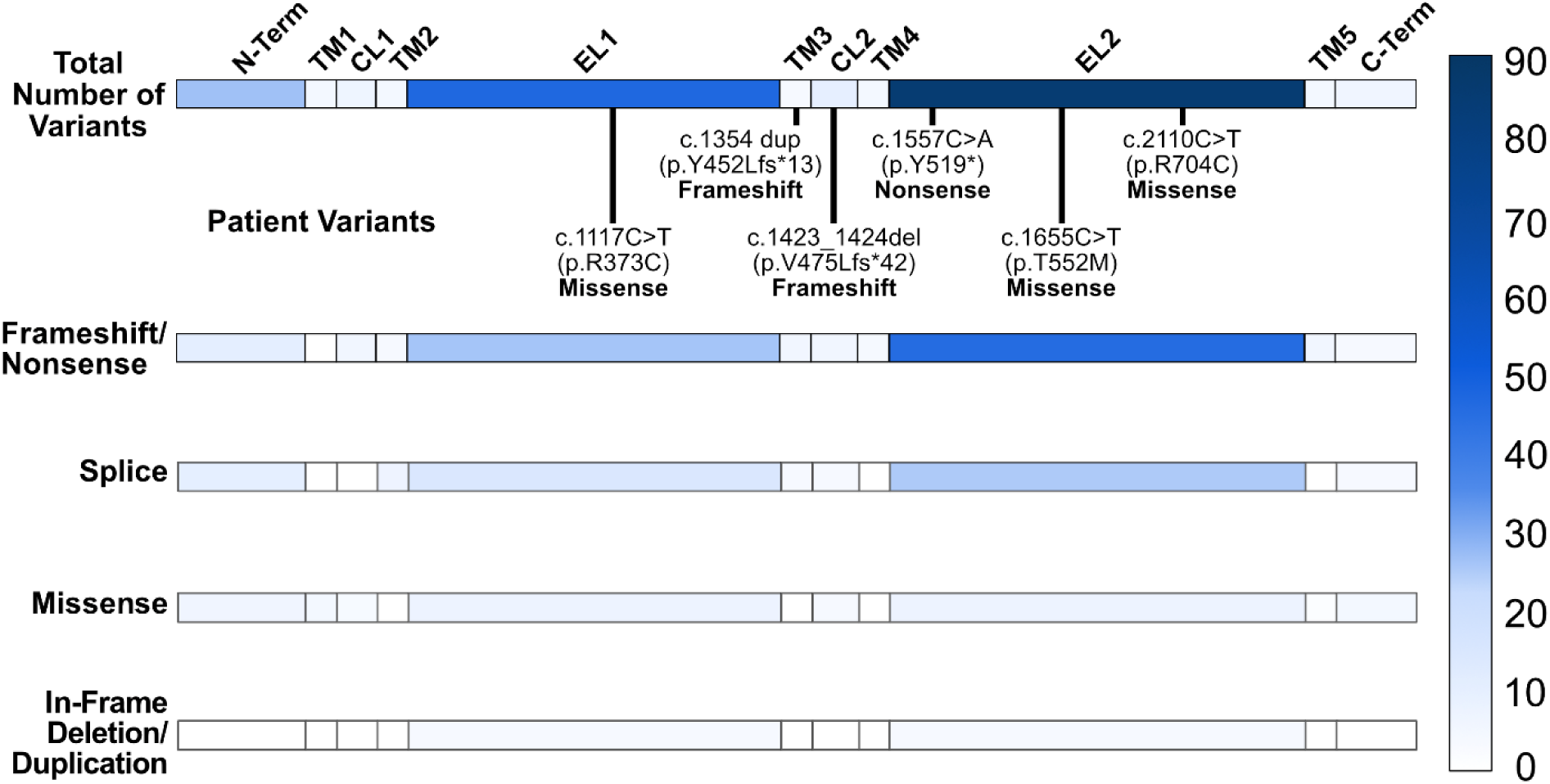
*PROM1* variant location in each protein domain, including the patients outlined in our case series. Most frameshift/nonsense, splice and missense variants occurred in extracellular loop 2, followed by extracellular loop 1, and the N-terminus. Specified variants represent the case series patients represented in this manuscript. Abbreviations: N-term, N-terminal; TM1-TM5, transmembrane domains 1-5; CL1-CL2; cytoplasmic loops 1 & 2; EL1-EL2, extracellular loops 1 & 2; C-Term, C-Terminal.

### Genotype-Phenotype-Inheritance Correlations

We identified two statistically significant associations from Monte Carlo chi-square analyses for genotype-phenotype-inheritance correlations: variant type vs. inheritance (x^2^ = 17.898, df = NA, p = 0.0005) and phenotype vs. variant type (x^2^ = 23.754, df = NA, p = 0.04848). For variance type vs. inheritance, standardized residuals revealed enrichment in observations of dominant inheritance of missense variants (z = 4.19); this relationship was identified to be strong by Cramér’s V analysis (V = 0.4257; **Fig. 4A**). For phenotype vs. variant type, standardized residual analysis revealed increased observations of missense variants associated with macular dystrophy (z = 3.41) and frameshift/nonsense variants associated with rod-cone dystrophy (z = 2.67); Cramér’s V identified these phenotype-variant relationships as having a moderate strength (V = 0.1585; **Fig. 4B**). Variant type vs. protein domain was not statistically significant (x^2^ = 35.865, df = NA, p = 0.2574) but standard residuals revealed an increase in observed cases of missense variants in transmembrane domain 1 (z = 3.97); this relationship was identified to have a weak strength by Cramér’s V (V = 0.1006; **Fig. 4C**). Inheritance vs. phenotype relationships did not achieve statistical significant (x^2^ = 5.5192, df = NA, p = 0.2909), but standardized residual analysis demonstrated enriched relationships between dominant inheritance of macular dystrophy (z = 2.30) (**Fig. 4D**); Cramér’s V identified this relationship as moderate (V = 0.1339). Regarding inheritance patterns and protein domains, there was no statistically significant relationship (x^2^ = 8.8516, df = NA, p = 0.4668), but an increase in observed cases of dominant inheritance in transmembrane domain 1 was detected by residual analysis (z = 2.14); Cramér’s V identified this relationship strength as negligible (V = 0.0; **Fig. 4E**). Finally, analysis of phenotype vs. protein domain relationships also did not reach statistical significance (x^2^ = 34.341, df = NA, p = 0.6767), and standardized residual analysis identified only slightly enriched observations of retinal dystrophy in cytoplasmic loop 1 (z = 2.13), rod-cone dystrophy in transmembrane domain 2 (z = 2.09), and Leber Congenital Amaurosis in extracellular loop 2 (z = 2.01); these relationships were also identified as negligible by Cramér’s V (V = 0.0, **Fig. 4F**).

**Figure 4.**
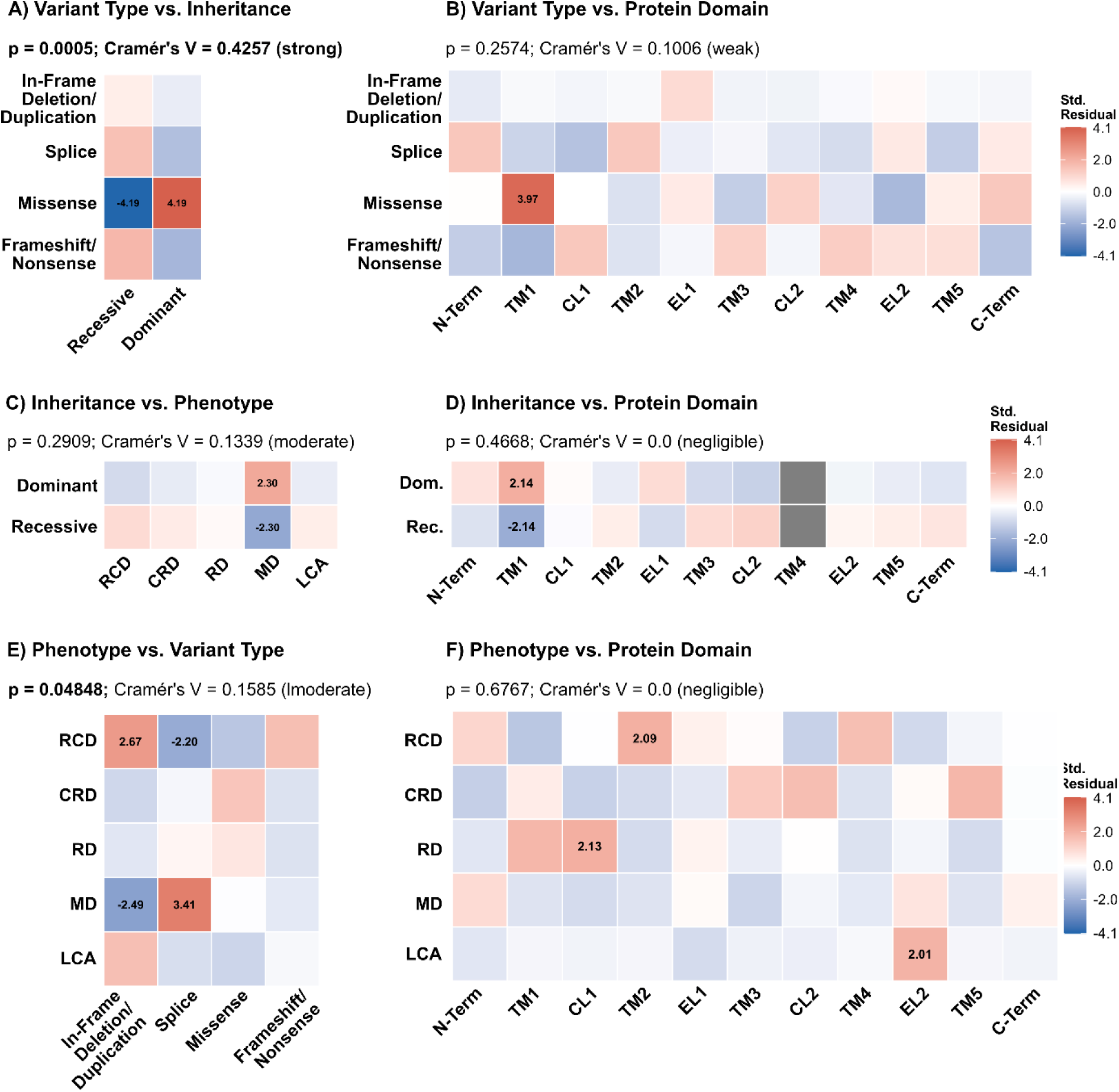
Heat maps representing standardized residuals from Monte Carlo chi-square analysis of *PROM1*-associated genotype-phenotype relationships. Heat maps represent variant type and inheritance (A), variant type and protein domain (B), inheritance vs. phenotype (C), inheritance vs. protein domain (D), phenotype vs. variant type (E), and phenotype vs. protein domain (F). Only variant type vs. inheritance achieved statistical significance (p = 0.0025, V = 0.44) and this was due to the association of dominant inheritance of missense variants. Other relationships of interest include missense variants associated with TM1, missense variant association with macular dystrophy, and frameshift/nonsense variant association with rod-cone dystrophy. Abbreviations: RCD, rod-cone dystrophy; CRD, cone-rod dystrophy; MD, macular dystrophy; RD, retinal dystrophy; LCA, Leber congenital amaurosis; N-term, n-terminal domain; TM3-TM5, transmembrane domains 3 and 5; CL2, cytoplasmic loop 2; EL1, extracellular loop 1; C-term, C-terminal domain.

### Case Series

Here, we report the clinical data from seven patients (P1-P7) with disease-causing *PROM1* variants. All patients in this series were unrelated, including P2 and P3 who had identical variants; two of the five patients were female. Patient demographics can be found in **Table 1**.

**Table 1.**
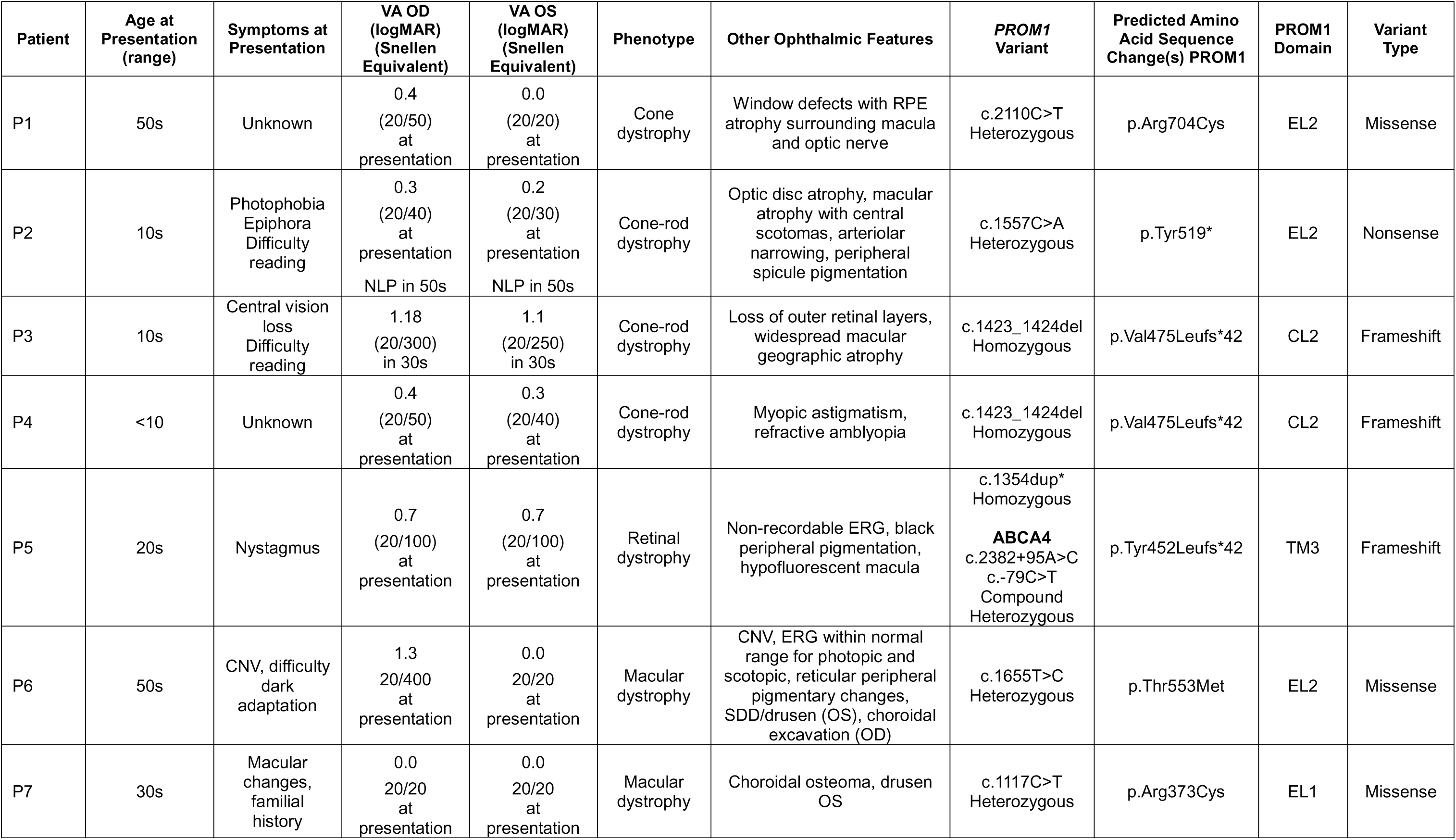
Patient demographics, ophthalmic features, genotype, and predicted amino acid change for *PROM1*-associated case series.

### Patient 1: Heterozygous c.2110C>T (p.Arg704Cys) pathogenic missense variant. Cone Dystrophy

Patient 1 has a heterozygous c.2110C>T, p.Arg704Cys missense variant. This patient presented with maculopathy in their 50s. Genetic testing was performed upon first presentation. Their visual acuity (VA) was OD 20/50 and OS 20/20. Fundus photos demonstrated macular atrophy (OD) and macular granularity and mottling (OS). Fundus autofluorescence (FAF) imaging showed corresponding areas of confluent macular hypoautofluorescence OD and parafoveal hypoautofluorescence and surrounding diffuse hyperautofluorescence of the macula OS (Fig. 5A-B). Visual field testing showed reduced macular sensitivity but preserved peripheral sensitivity in both eyes (Fig. 5C-D). Photopic full-field ERG with 3.0 cd s/m2 flash intensity demonstrated a significant reduction in cone a-wave and b-wave responses (OD: a-wave 9.8 µV, b-wave 34.3 µV; OS: a-wave 18.4 µV, b-wave 28.2 µV; Fig. 5E) indicating impairment of cone function. Scotopic ERG for the pure rod response was within normal ranges (OD: b-wave 96.6 µV; OS: b-wave 104.7 µV; Fig. 5F). Given the clinical fundus and FAF findings, and the ERG demonstrating impairment of cone responses with a pure rod response within normal limits, this patient’s phenotype was classified as cone dystrophy. This variant was classified as pathogenic and represents a rare cause of *PROM1*-related retinal dystrophy^27^. The phenotype of this patient correlates well with our statistical data, which indicate a strong relationship between missense variants and a dominant form of milder macular or cone-specific dystrophies.

**Figure 5.**
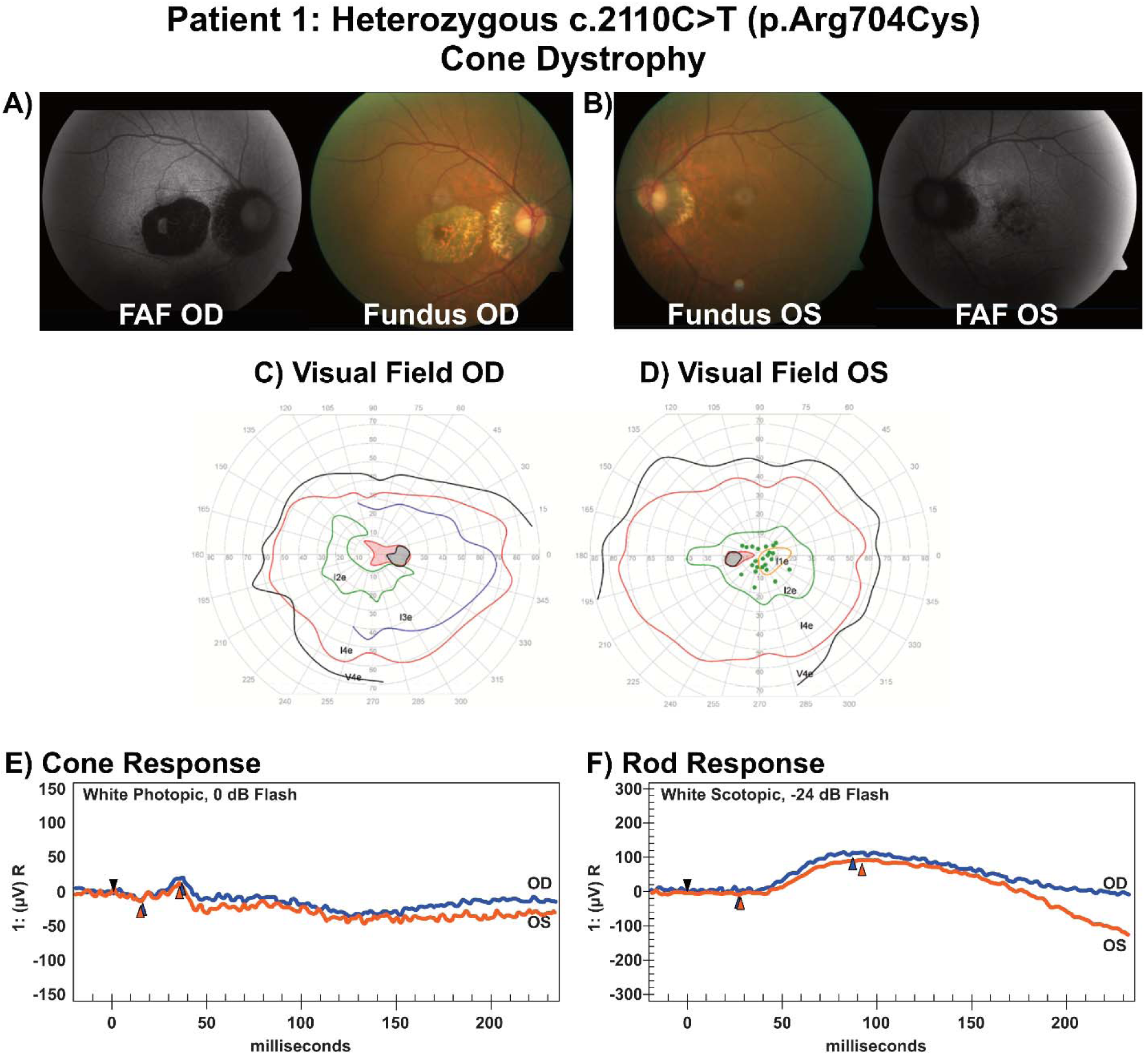
Clinical data from Patient 1, who has a pathogenic heterozygous c.2110C>T missense variant and clinical findings that support a diagnosis of cone dystrophy. A-B) represent color fundus and fundus autofluorescence photos with a primary finding of hypofluorescence surrounding the macula and optic nerve. C-D) Visual field data for OD and OS. OS is more severely affected, with several non-seeing areas surrounding the macula (green dots). E-F) Photopic (cone) ERG responses are nearly non-existent whereas the scotopic ERG (pure rod response) is within normal limits (OD, blue; OS, orange).

### Patient 2: Homozygous c.1557C>A (p.Tyr519*) pathogenic variant. Cone-Rod Dystrophy

Patient 2 has a long natural history in our clinic; we have clinical natural history of vision loss beginning in childhood (5-10 yrs), and they are now in their 50s. Fundus photos in early disease stages showed primarily macular disruptions, but as the retinal degeneration progressed, greater peripheral effects were seen, including bone spicules and increased hypoautofluorescence in the peripheral retina (**Fig. 6A-B**). OCT scans showed increased disruption of the photoreceptor layer and foveal thinning as the patient aged (**Fig. 6C-D**). In their teens, their VA was OD 20/40 and OS 20/30, progressing to 20/100 and 20/80 in their late 20s, hand motion (HM) OU in their 30s, and no light perception (NLP) in their 40s. During the time this patient was followed, they have experienced progressive loss of photoreceptors, significant central macular thinning, central scotomata, and greater disruption of cone ERG responses, compared to rod responses (**Fig. 6E**). This patient’s diagnosis is “cone-rod dystrophy”, which matches the database classification for this variant.

**Figure 6.**
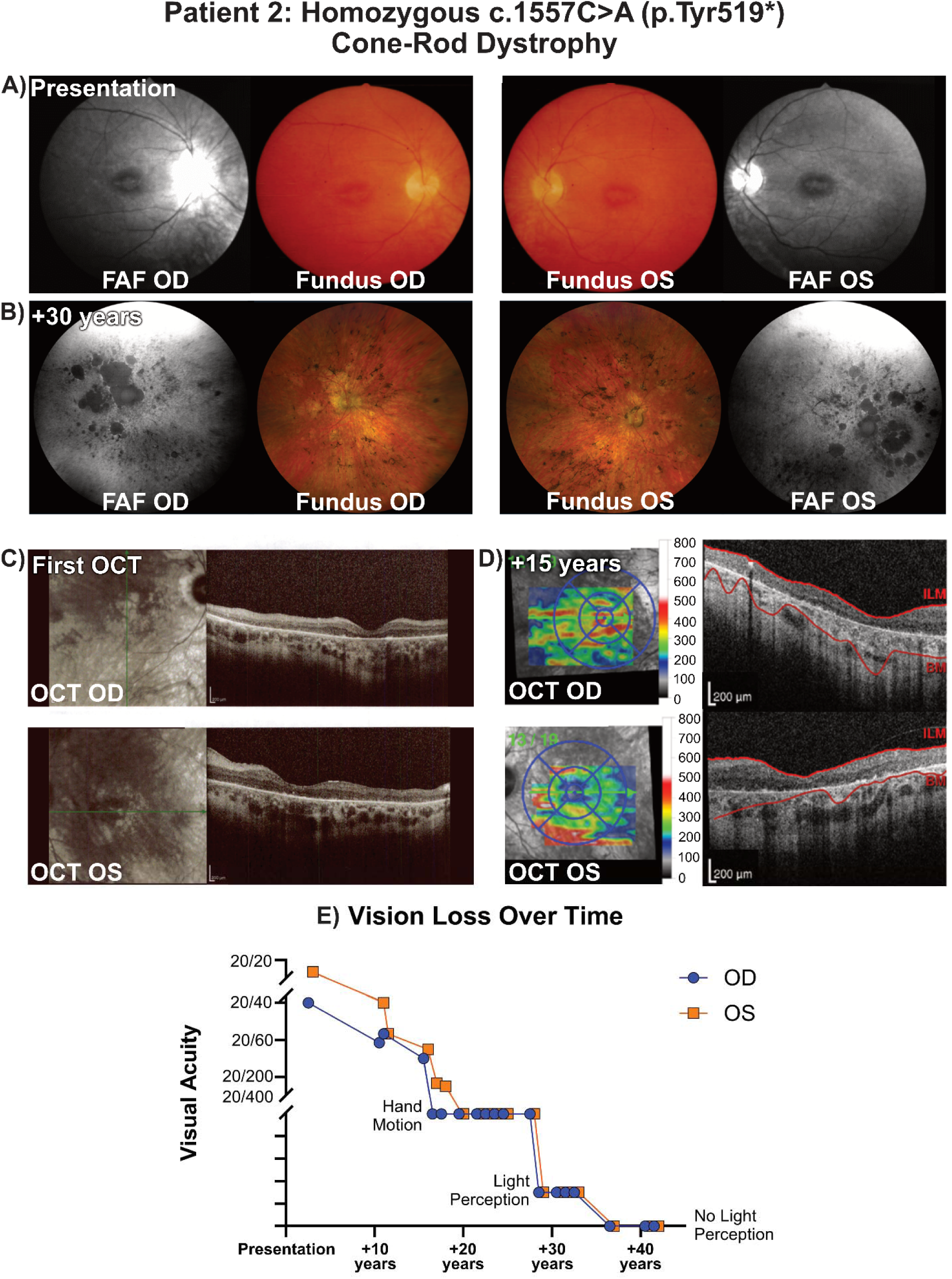
Clinical data from patient 2, who has a homozygous c.1557C>A nonsense variant, cone-rod dystrophy, and 40+ years of clinical follow up. A-B) Fundus and red-free photos demonstrate hypopigmentation and blunted macular reflex on color photos and evidence of increased foveal attenuation on red-free fundus images. (A) to a broader cone-rod dystrophy phenotype, including the increased presence of bone spicule-like pigmentary changes and increased hypoautofluorescent lesions in the periphery (B). C-D) OCT changes demonstrate foveal thinning and loss of photoreceptors over time. E) Visual acuity loss progressed from 20/20 to 20/40 to hand motion in the span of 20 years, and then progressed from hand motion to no light perception in another 15 years (OD, blue; OS, orange).

### Patients 3 and 4: Homozygous c.1423_1424del (p.Val475Leufs*42) pathogenic variant

Patients 3 and 4 are unrelated, but both have the same homozygous c.1423_1424del, p.Val475Leufs*42 variant. Patient 3 first presented in their teens (10-15). Fundus photos were taken and this patient was given a diagnosis of cone-rod dystrophy; there was little progression between teenage years into their 20s (**Fig. 7A,B**). By their 30s, the patient complained of difficulty reading, and their VA was recorded as OD 20/300 and OS 20/250. Fundus photos and FAF demonstrated RPE mottling, temporal optic nerve pallor, and significant macular hypoautofluorescence (**Fig. 7C)**. The diagnosis of cone-rod dystrophy is different from the variant classification according to online databases (macular dystrophy), but it does fall in line with our correlation analysis, which indicates that recessive variants are more likely to cause pan-retinal dystrophies instead of macular- or cone-specific phenotypes.

**Figure 7.**
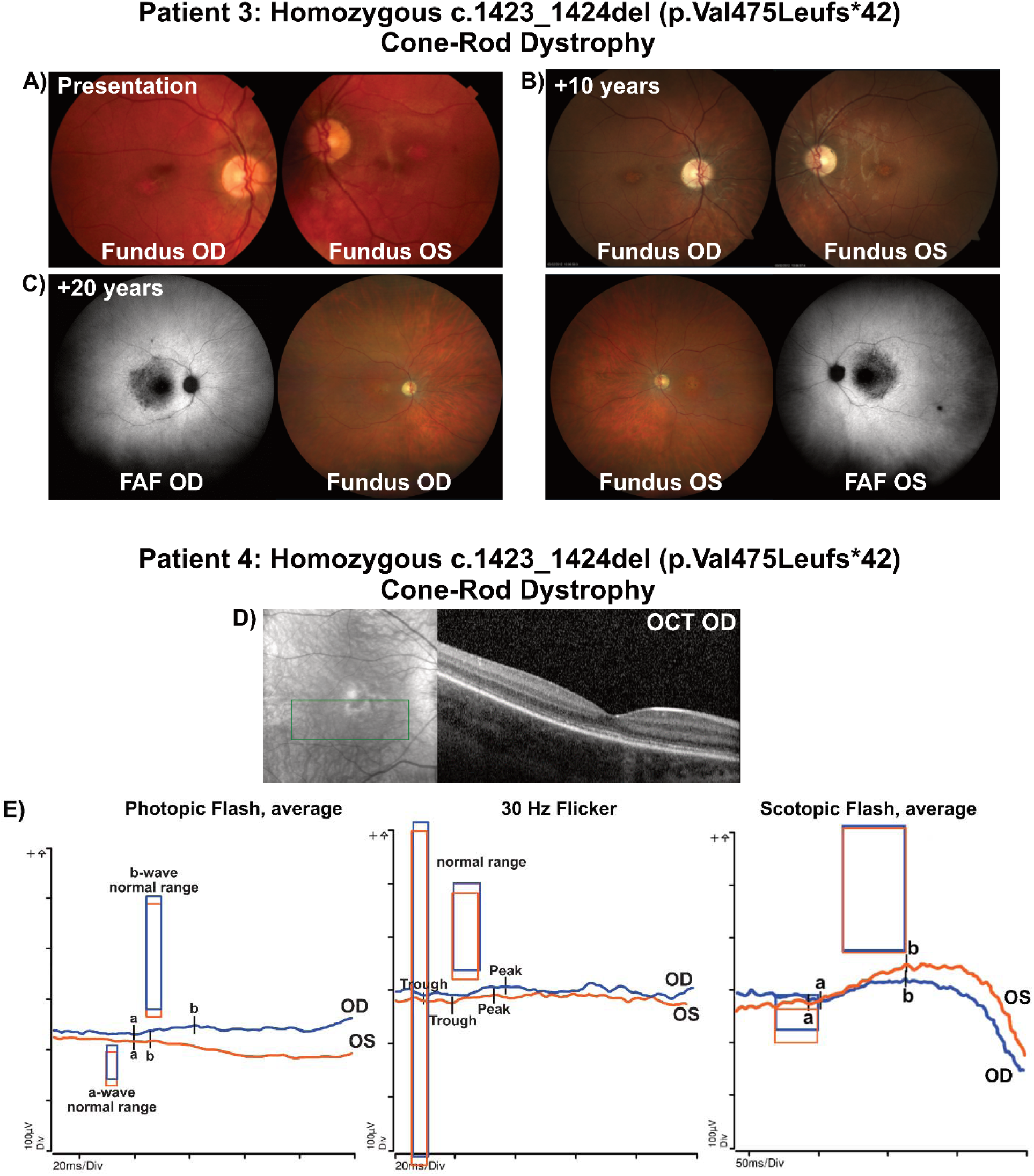
Clinical data for two patients with a homozygous c.1423_1424del, p.Val457Leufs*42 pathogenic frameshift variant who were both diagnosed with cone-rod dystrophy. A-C) Fundus photos from patient 3, macular pigmentary changes and FAF hypoautofluorescence. D) Patient 4’s OD OCT scan demonstrates ellipsoid zone disorganization and macular changes. E) Patient 4’s ERG data support a diagnosis of cone-rod dystrophy, due to significantly greater impairment of the cone response compared to the rod response (OD, blue; OS, orange).

Patient 4 presented in early childhood (< 5 years old). Their VA was OD 20/50 and OS 20/40, and they were identified as having myopic astigmatism and refractive amblyopia. An undilated fundus exam revealed healthy appearing optic discs, vessels, and maculae bilaterally, but OCT revealed ellipsoid zone disorganization and loss (**Fig. 7D)**. Full-field ERG from both eyes had reduced b-wave amplitudes for scotopic 0.01 cd s/m2 flash, photopic 3.0 cd s/m2 flash, and photopic 30 Hz flicker series. Photopic a- and b-waves were more significantly reduced than scotopic b-waves, so this patient was diagnosed as having cone-rod dystrophy (**Fig. 7E**). This classification matches patient 3, and expectations from our statistical analysis.

### Patient 5: Homozygous *PROM1* c.1354dup, p.Y452Leufs*13 pathogenic variant

Patient 5 presented in their 20s. Their VA was recorded as OD 20/100 and OS 20/100. Fundus photographs showed blunted macular reflexes and foveal atrophy as well as RPE granularity and pigmentation in the mid-periphery (**Fig. 8A**). FAF demonstrated foveal hypoautofluorescence and speckled hyper- and hypo-fluorescence in the surrounding region (**Fig. 8B**). OCT scans demonstrate preservation of the RPE, but significant loss of the ellipsoid zone and thinning of the ONL indicating photoreceptor loss; there was some preservation of the sub-foveal ellipsoid zone OU (**Fig. 8C-D**). The patient’s scotopic and photopic ERGs did not produce any measurable responses; thus, their phenotype was classified as severe retinal dystrophy (**Fig. 8E-F**). This does not match the database classification of cone-rod dystrophy for this variant; however, it is possible that we would have uncovered a cone-rod dystrophy pattern in this patient had a full field ERG been performed at an earlier age. In addition, the patient had compound heterozygous VUSs in *ABCA4*: c.2382+95A>C and c.-79C>T, a gene associated with retinal disease that can phenocopy *PROM1*-related disease. While these variants are unlikely to be the primary driver of the patient’s retinal disease, it is difficult to completely exclude a potential modifier effect.

**Figure 8.**
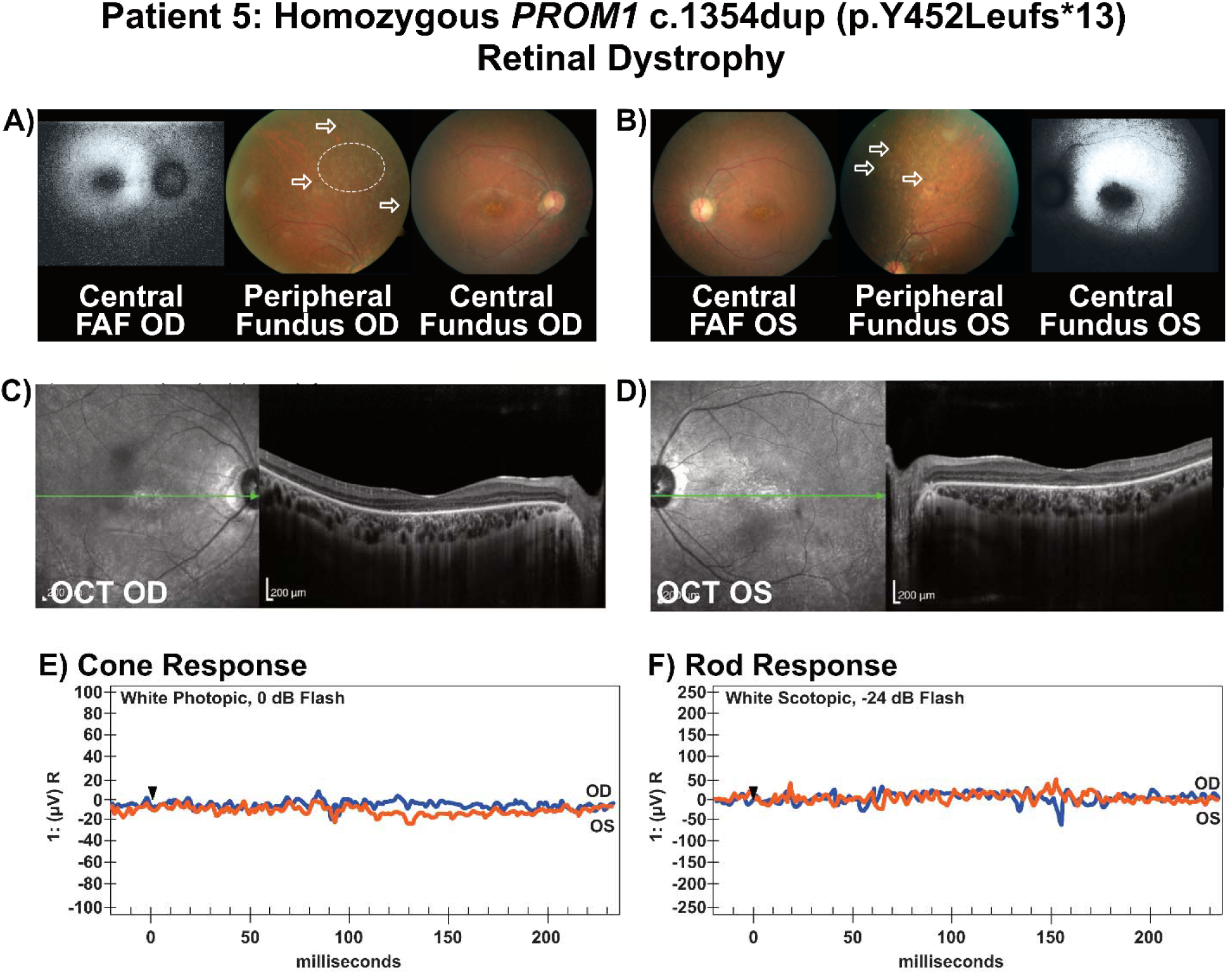
Clinical data for Patient 5, who has a homozygous c.1354dup pathogenic frameshift variant, that support the finding of retinal dystrophy. A-B) Fundus photography and FAF revealed both macular and peripheral abnormalities including macular hypopigmentation and hypofluorescence, RPE granularity in the mid-periphery (OD, white circle), and abnormal hyperpigmentation in the mid-periphery (OU, white arrows). C-D) OCT scans revealed foveal thinning, ellipsoid thinning, and loss of the ONL. E-F) Photopic and scotopic ERG responses were undetectable for both eyes (OD, blue; OS, orange).

### Patient 6: Heterozygous c.1655C>T (p.Thr553Met) missense VUS. Macular Dystrophy

Patient 6 presented in their 50s with no known prior family history of retinal disease and difficulty with dark adaptation. Their VA was recorded as OD 20/400 and OS 20/20. Fundus photos show central macular atrophy with pigmentary changes and an appearance of inactive CNV. There are also a few surrounding drusen. The peripheral fundus has reticular pigmentary changes (**Fig 9A**). OS showed macular drusen, pigmentary changes, and an appearance of inactive CNV, with a small flat choroidal nevus temporal to the fovea; the peripheral fundus also has reticular pigmentary changes (**Fig. 9B**). The optic nerves look normal in both eyes. OD OCT revealed severe macular atrophy with loss of outer retinal lamination, as well as a thickened choroid with pronounced subfoveal choroidal excavation. Confluent drusen are evident at the edges of the atrophy (**Fig. 9C)**. For OS OCT, there are numerous drusen and subretinal drusenoid deposits, with a few areas of overlying ellipsoid zone disruption; the choroid also appears thickened (**Fig. 9D**). Photopic and scotopic ERG amplitudes fall within normal limits (**Fig. 9E-F**). This patient was classified as having macular dystrophy with peripheral retinal pigmentary changes. To the best of our knowledge, this variant has not yet been described in the medical literature, but that there is a single entry in ClinVar (ID 1914531; clinical indication for genetic testing was not reported).

**Figure 9.**
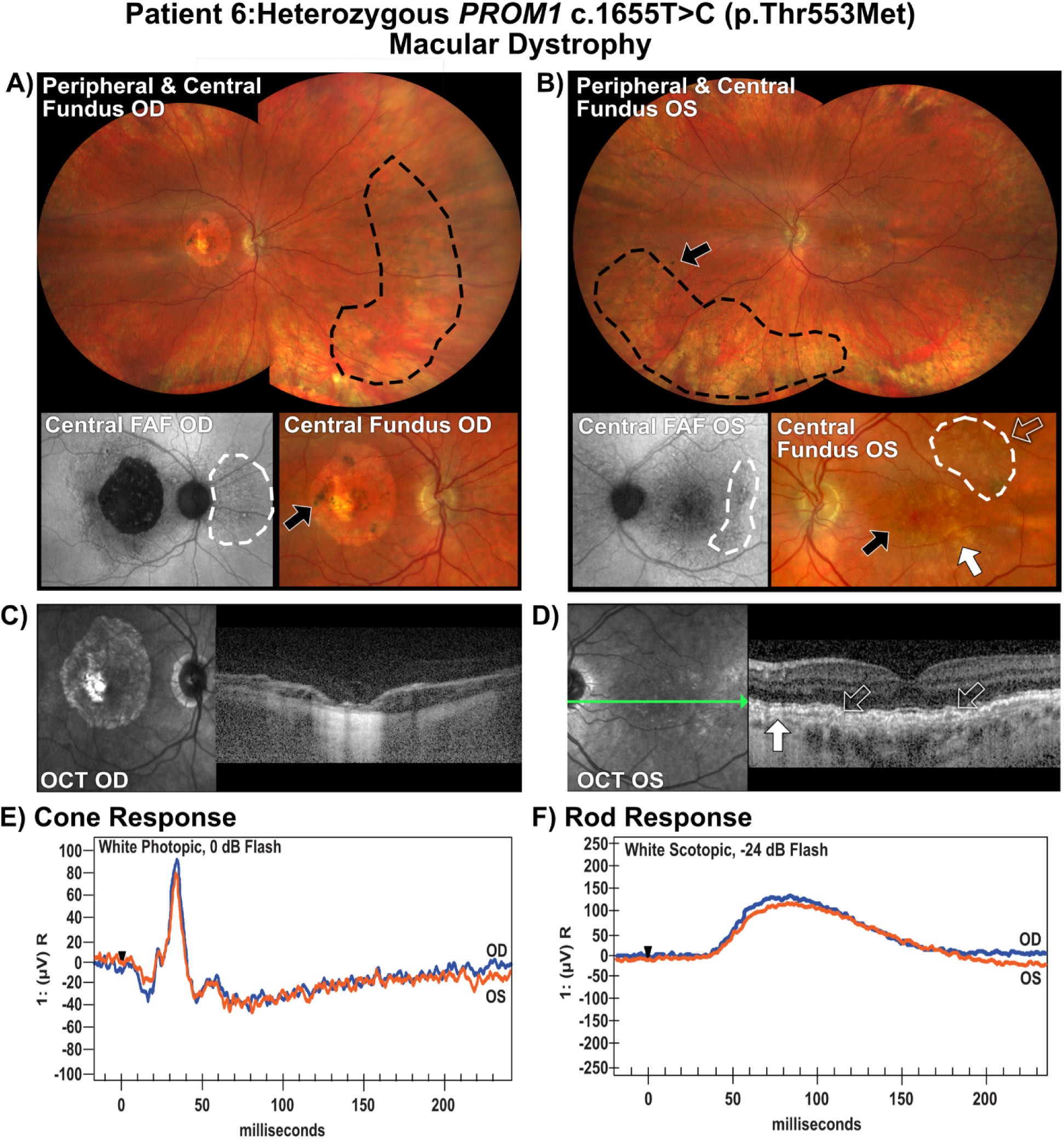
Clinical data for Patient 6, who has a heterozygous p.Thr553Met pathogenic missense variant, that support the finding of macular dystrophy. A-B) Fundus photography and FAF revealed both macular and peripheral abnormalities including macular hypopigmentation, peripheral pigmentary changes (black outlines, black arrows), SDD (white outlines, clear arrows), and soft drusen (white arrows). C-D) OCT scans revealed severe retinal atrophy and choroidal excavation in OD and SDD (clear arrows) and soft drusen (white arrows) in OS. E-F) Photopic and scotopic ERG responses were detectable, but dampened for both eyes (OD, blue; OS, orange).

### Patient 7: Heterozygous c.1117C>T (p.Arg373Cys) pathogenic missense variant. Macular Dystrophy

Patient 7 presented in their late 30s with a known prior family history of retinal disease. Their VA was recorded as OD 20/20 and OS 20/20. Fundus photos showed foveal mottling (OU) and choroidal osteoma (OS). FAF showed macular hypoautofluorescence surrounded by speckled hyperfluorescence OU (**Fig. 10A, B**). OCT scans showed parafoveal thinning and an accumulation of hyperreflective material under the fovea in OU (**Fig. 10C,D**); OS also had drusenoid hyperreflective deposits (**Fig. 10D**). On the basis of these clinical data, this patient’s phenotype was classified as macular dystrophy, which is typical of the p.Arg373Cys pathogenic variant.

**Figure 10.**
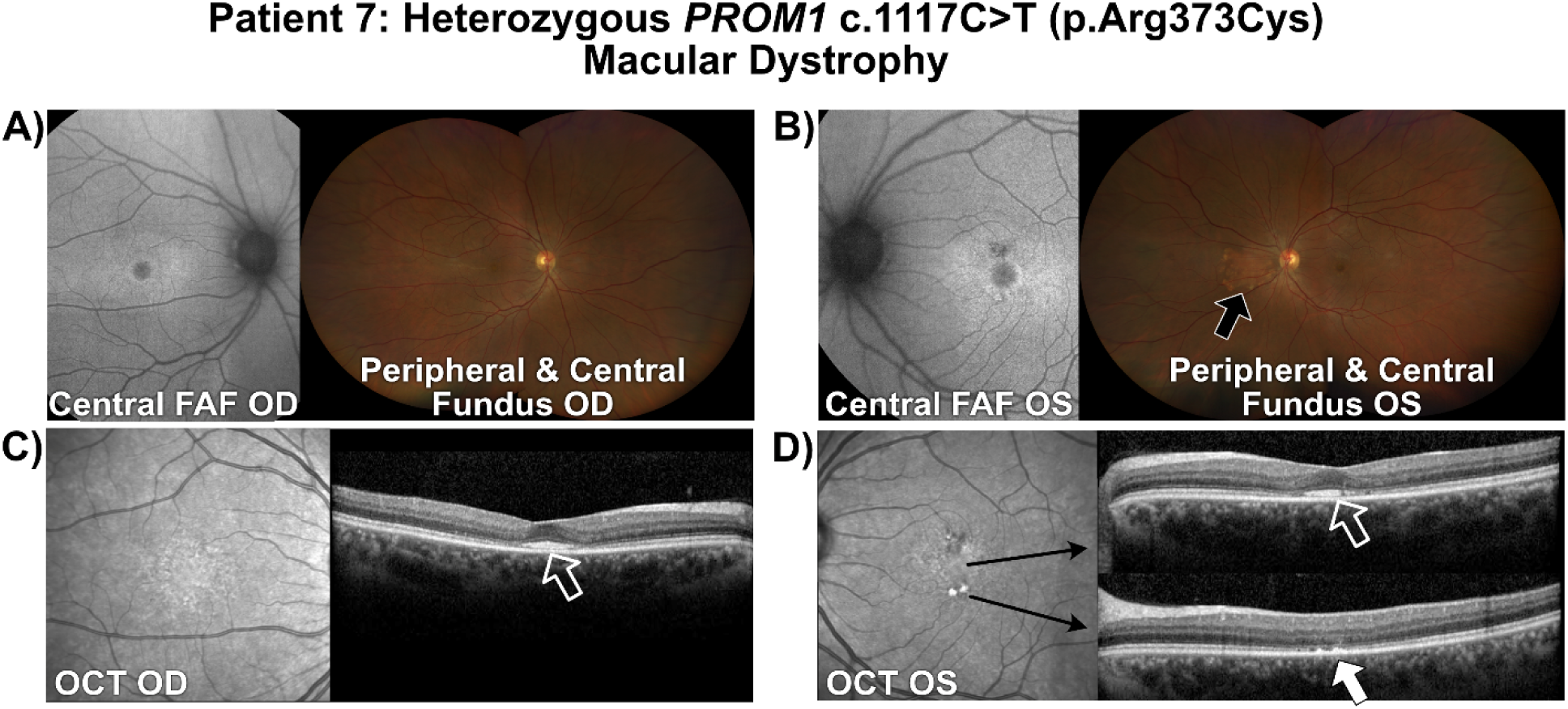
Clinical data for Patient 7, who has a heterozygous p.Arg373Cys pathogenic missense variant, that support the finding of macular dystrophy. A-B) Fundus photography and FAF reveal foveal mottling and FAF changes, and an incidental choroidal osteoma OS (black arrow). C-D) OCT scans revealed parafoveal thinning and build up of hyperreflective material under the fovea (clear arrows), and drusenoid hyperreflective deposits (white arrows) in OS.

## Discussion

In this study, we formally analyzed genotype-inheritance-phenotype relationships for 190 reported pathogenic or likely pathogenic *PROM1* variants found in two genetic databases: ClinVar and HGMD. We found a strong, statistically significant association between dominant inheritance of missense variants, and strong associations between missense variants and macular or cone-specific dystrophies. These findings confirm a previous report of associations of missense variants with mild macular dystrophy, which was intuitively determined after examining a cohort of 19 patients with *PROM1-*associtaed retinal degeneration^3^. Truncating frameshift/nonsense variants, on the other hand, are generally recessive and associated with more severe forms of pan-retinal degeneration such as cone-rod dystrophy, retinal dystrophy, and retinitis pigmentosa^3,–7,13,32^; our statistical data support this conclusion as well.

Missense variants can be benign or harmful, and mechanisms associated with the pathogenesis of missense variants often correlate with changes in the three-dimensional structure of proteins, which depend on amino acid charge and interactions to maintain their structure^29,30^. This can cause mild to severe effects on the protein and therefore cellular health, ranging from mild functional impairment and loss of binding to functional partners to toxic protein aggregation and cell death^31^. It can also lead to weakening of cellular scaffolding due to alterations in protein binding, leading to structural instability of the cell or cell membrane^32^. This scaffolding weakening effect could be particularly important for PROM1, due to its role as a lipid raft organizer its involvement in photoreceptor outer segment morphogenesis and maintenance^5(p2021),12,13,19,20^. It is potentially the case that the correlation with missense variants and macular dystrophy is due to a partially functional protein, instead of complete loss or a dominant negative effect. Rods have the extra support of the plasma membrane and disc fusion to aid with outer segment integrity, whereas cones lack disc fusion and an outer plasma membrane, and thus have less structural support, making them less able to cope with even mild impairments in PROM1 function, leading to a more cone-specific macular dystrophy type phenotype.

Frameshift/nonsense variants are associated with an increased risk of pathogenicity due to their ability to dramatically alter the protein structure or prevent protein synthesis altogether^33^. Complete loss of *PROM1* through improper splicing, nonsense-mediated mRNA decay, or truncating frameshifts results in severe rod and cone photoreceptor outer segment disorganization in animal models, which likely corresponds to more severe pan-retinal dystrophies^12,13,19,34–36^. Splice site variants disrupt protein function typically by exon skipping, but they can also activate cryptic splice sites and cause pseudoexon and/or intron retention, usually leading to a premature stop codon. *PROM1* has multiple splice variants in the N-terminus, extracellular loop 1, and the C-terminus which have tissue-specific localization within the body^37,38^. In humans, this alternate splicing gives rise to seven different protein isoforms – s1, s2, s7, s9, s10, s11, and s12 – which are expressed differentially in specific organ and tissue systems^38^. Isoforms s11 and s12 are expressed highly in the retina, whereas isoform s2 (canonical PROM1) is represented at a much lower level. Phenotypic heterogeneity could also reflect yet unknown subtle variations in the function of the PROM1 protein in rods versus cones; this is an under-studied area of research.

There was a significant preference for disease causing *PROM1* variants to be localized to the extracellular loops (66% of total variants). The most likely explanation for this is that these loops also represent nearly 2/3 of the total protein amino acids (∼62% of total amino acids), so it is simply a proportional representation of variants within *PROM1*. In support of this, there were no statistically significant or enriched relationships regarding EL1 or EL2 protein domains in our analysis; LCA-associated variants were all contained within EL2, however, which did lead to a small increase in observed vs expected values for this phenotype (z = 2.01). Furthermore, analysis of benign *PROM1* missense variants also represent approximately 2/3 of the total protein (5/9 = 56% identified variants from gnomAD – accessed July 2026).

The structure of PROM1 is tightly regulated through post-translational modifications and therefore subtle changes can influence its function. PROM1 has nine extracellular N-linked glycosylation sites, and highly conserved cysteine, lysine, and tyrosine residues that contribute to protein folding, stability, membrane localization, and post-translational regulation^39–41^. Important PROM1 motifs in the extracellular loops that could impact its function include lysine residues in EL1 at p.K216, p.K248, and p.K255, that are acetylated to stabilize PROM1 as it is transported to the cell membrane^40^; if the acetylation is disrupted, prominin becomes unstable and cell surface localization is impaired, which could then affect PROM1 function. The extracellular loops also contain nine N-glycosylation sites. N-glycosylation is a posttranslational modification that can impact protein function and stability, and thus alterations in glycosylation could contribute to the extracellular loops being a hotspot for disease causing variants^40^. Outside of the retina, loss of single glycosylation sites does not appear to significantly affect PROM1 expression or membrane localization for most sites^40^, but impaired glycosylation at p.Asn548 in EL2 was reported to affect interactions between PROM1 and beta-catenin, inhibiting PROM1-mediated cell proliferation^39^. The effects of modified protein glycosylation in retinal PROM1 remain largely unknown. It has been demonstrated that the retina has specific PROM1 glycosylation patterns compared to other cell types, however, and it was suggested that this may aid PROM1 in its unique role in photoreceptor outer segment membrane organization^42^.

Other potentially interesting, but not statistically significant, correlations related to protein domains include a large increase in observed counts of missense variants in transmembrane domain 1 (z = 3.97) which are likely to be dominantly inherited (z = 2.14), retinal dystrophy associated with cytoplasmic loop 1 (z = 2.13), and rod-cone dystrophy associated with transmembrane domain 2 (z = 2.09). Transmembrane domain 1 has a large cluster of cysteine residues, which play a role in protein folding and scaffolding through the linking of covalent disulfide bonds^43,44^. PROM1 is closely associated with cholesterol containing lipid rafts^14,18,45–47^, and cysteine residues can also play a significant role in targeting proteins to sphingolipid and cholesterol rich lipid membrane rafts through palmitoylation^40^. Therefore, modification of this site could disrupt how PROM1 is trafficked to, or interacts with, the lipid membrane or other cell-surface proteins, creating a dominant disease-associated variant hotspot near TM1 and CL1^40^. There is also a missense variant associated with cone-rod dystrophy (p.Trp795Arg) in TM5 that has been reported to cause increased binding of cholesterol in stably transfected Expi293 cells^47^. Changes in how PROM1 interacts with cholesterol and therefore impacts membrane curvature could preferentially affect rods versus cones. There are no known significant protein domains or clusters in TM2; the small enrichment in observed values is most likely because reported variants in TM2 are so far associated only with rod-cone dystrophy.

Our correlation analysis revealed potential genotype-phenotype relationships for only a small proportion of the total *PROM1* variants, so we must look elsewhere to explain the large differences in clinical manifestations reported for *PROM1*-associated disease. Other sources of heterogeneity could be individual differences in variant penetrance and expressivity, genotype-environment interactions, and difficulties in accurate clinical diagnosis in patients seen at a single time point or with missing data. Studies on genetic background have shown that the behavior of a variant in different individuals can be influenced by the presence of a large number of loci on different alleles, altering the variant expression in one individual compared to the expression of the same variant in another individual^48–50^. Variable expressivity is particularly common in dominantly inherited conditions. The penetrance and expressivity of a variant can also change due to gender- or age-related factors, epigenetics, genetic modifiers, and environmental factors (diet, alcohol, drugs, and metabolic syndromes)^51^. Patient 5 is a potential example of clinical heterogeneity caused by genetic background; they have a *PROM1* variant that is reported in databases to be associated with cone-rod dystrophy (c.1354dup), but instead demonstrate a severe retinal dystrophy. This patient also has compound heterozygous VUS for *ABCA4* (c.2382+95A>C, c.-79C>T), a gene associated with Stargardt macular dystrophy, and it has been reported previously that modification in *ABCA4* can compound the effects of *PROM1*-associated blindness^52^. Thus, genetic interactions between *PROM1* and *ABCA4* (or other unknown genes) could contribute to the difference between the reported phenotype for this variant and our patient phenotype, and other patients in the literature. The complex relationships between individual factors and penetrance and expressivity remain largely unknown, and are areas that would benefit from further research^50^.

Another potential source of reported heterogeneity for *PROM1*-associated retinal degenerations is differences in clinical diagnoses, or diagnoses that may change over time as retinal degeneration progresses and/or new information is gained. Clinical diagnoses of retinal disease are based on the information at hand, and it can be difficult to definitively assign a phenotype when there are missing data or missing information about the patient’s natural or familial history. For example, it can be difficult to obtain fundus photos, OCT, or ERG in young children or persons with significant blindness or nystagmus, as lack of fixation or excessive movement can significantly alter or void the results of these types of exams. There may also be differences in access to these types of equipment or to specialist care by an ophthalmologist, leading to gaps in the patient’s natural history. Diagnoses made without having access to the full natural history of a patient may also not tell the entire story, and it is possible that diagnosis of phenotypes may change over time as new information is gained.

Finally, the use of AI and deep-learning genetic algorithms to mine genetic data and predict variant outcomes is a powerful tool that is becoming more common. The use of algorithms can help to identify potential pathogenicity of VUS without patient data, but may lead confusion if incorrect outcomes are reported in databases and it is not specified or noticed that they are predictions and not verified with clinical data.

In conclusion, the source of PROM1-associated disease heterogeneity, whether associated with protein structure, protein function in rods versus cones, or differential diagnoses of clinical phenotypes, remains mostly enigmatic. Given the small sample size, however, we were able to determine statistically significant positive associations between missense variants and dominantly inherited macular dystrophy and frameshift/nonsense variants with recessive inheritance of more severe retinal dystrophies that include rod and cone dysfunction. While these results should be interpreted with caution, they may provide predictive value regarding future *PROM1*-associated disease expectations that could be used by clinicians and patients to guide strategies for patient visits and follow up. The underlying causes of clinical heterogeneity of monogenic disease is a question that plagues many other forms of inherited diseases. Understanding further potential reasons why this variance exists would aid in more efficient and accurate diagnosis, and better study design for future therapeutics to treat *PROM1*-associated retinal degenerative disease.

## Data Availability

All data produced in the present work are contained in the manuscript.

