## Supplemental Tables S2-7 for "Genotype-Phenotype Correlations Reveal Positive Inheritance and Phenotype Associations for *PROM1*-Associated Inherited Retinal Degenerations"

**Table S2. Inheritance pattern vs. variant type chi-square analysis.** Increased observed counts are highlighted in red, and decrease counts are shown in blue.

| **Inheritance vs Variant Type Observed** | | | | |
|---|---|---|---|---|
| | Frameshift | Missense | Splice | Deletion |
| Dominant | 5 | 9 | 1 | 0 |
| Recessive | 41 | 8 | 18 | 1 |

| **Inheritance vs Variant Type Expected** | | | | |
|---|---|---|---|---|
| | Frameshift | Missense | Splice | Deletion |
| Dominant | 8 | 3 | 3 | 0 |
| Recessive | 38 | 14 | 16 | 1 |

Monte Carlo Chi-Square
x-squared = 17.898, df = NA, **p-value = 0.0005**

| **Inheritance vs Variant Type Residuals** | | | | |
|---|---|---|---|---|
| | Frameshift | Missense | Splice | Deletion |
| Dominant | -1.90 | **4.19** | -1.65 | -0.47 |
| Recessive | 1.90 | **-4.19** | 1.65 | 0.47 |

**Cramér's V** = 0.4257 *Strong

**Table S3. Inheritance pattern vs. phenotype chi-square analysis.**

| Inheritance vs Phenotype Observed | | | | | |
|---|---|---|---|---|---|
| | RCD | CRD | RD | MD | LCA |
| Dominant | 4 | 3 | 3 | 5 | 0 |
| Recessive | 27 | 18 | 15 | 7 | 1 |

| Inheritance vs Phenotype Expected | | | | | |
|---|---|---|---|---|---|
| | RCD | CRD | RD | MD | LCA |
| Dominant | 6 | 4 | 3 | 2 | 0 |
| Recessive | 25 | 17 | 15 | 10 | 1 |

Monte Carlo Chi-Square
x-squared = 5.5192, df = NA, p-value = 0.2909

| Inheritance vs Phenotype Residuals | | | | | |
|---|---|---|---|---|---|
| | RCD | CRD | RD | MD | LCA |
| Dominant | -0.94 | -0.52 | -0.18 | **2.30** | -0.47 |
| Recessive | 0.94 | 0.52 | 0.18 | **-2.30** | 0.47 |

**Cramér's V** = 0.1339 *Moderate

**Table S4. Inheritance pattern vs. protein domain chi-square analysis.** Increased observed counts are highlighted in red, and decrease counts are shown in blue.

| **Inheritance vs. Protein Domain Observed** | | | | | | | | | | | |
|---|---|---|---|---|---|---|---|---|---|---|---|
| | N-Term | TM1 | CL1 | TM2 | EL1 | TM3 | CL2 | TM4 | EL2 | TM5 | C-Term |
| Dominant | 2 | 1 | 1 | 0 | 5 | 0 | 0 | 0 | 6 | 0 | 0 |
| Recessive | 5 | 0 | 4 | 1 | 15 | 4 | 6 | 0 | 30 | 1 | 2 |

| **Inheritance vs. Protein Domain Expected** | | | | | | | | | | | |
|---|---|---|---|---|---|---|---|---|---|---|---|
| | N-Term | TM1 | CL1 | TM2 | EL1 | TM3 | CL2 | TM4 | EL2 | TM5 | C-Term |
| Dominant | 1 | 0 | 1 | 0 | 4 | 1 | 1 | 0 | 7 | 0 | 0 |
| Recessive | 6 | 1 | 4 | 1 | 16 | 3 | 5 | 0 | 29 | 1 | 2 |

Monte Carlo Chi-Square
x-squared = 8.8516, df = NA, p-value = 0.4668

| **Inheritance vs. Protein Domain Residuals** | | | | | | | | | | |
|---|---|---|---|---|---|---|---|---|---|---|
| | N-Term | TM1 | CL1 | TM2 | EL1 | TM3 | CL2 | EL2 | TM5 | C-Term |
| Dominant | 0.75 | **2.14** | 0.12 | -0.47 | 0.92 | -0.96 | -1.19 | -0.29 | -0.47 | -0.67 |
| Recessive | -0.75 | **-2.14** | -0.12 | 0.47 | -0.92 | 0.96 | 1.19 | 0.29 | 0.47 | 0.67 |

*TM4 removed from chi-square analysis

**Cramér's V** = 0.0 *Negligible

**Table S5. Phenotype vs. variant type chi-square analysis.** Increased observed counts are highlighted in red, and decrease counts are shown in blue.

| **Phenotype vs. Variant Type Observed** | | | | |
|---|---|---|---|---|
| | Frameshift | Missense | Splice | Deletion |
| RCD | 42 | 7 | 14 | 2 |
| CRD | 15 | 6 | 13 | 0 |
| RD | 16 | 7 | 11 | 0 |
| MD | 6 | 10 | 6 | 0 |
| LCA | 3 | 0 | 0 | 0 |

| **Phenotype vs. Variant Type Expected** | | | | |
|---|---|---|---|---|
| | Frameshift | Missense | Splice | Deletion |
| RCD | 34 | 12 | 18 | 1 |
| CRD | 18 | 6 | 9 | 0 |
| RD | 18 | 6 | 9 | 0 |
| MD | 11 | 4 | 6 | 0 |
| LCA | 2 | 1 | 1 | 0 |

Monte Carlo Chi-Square
x-squared = 23.754, df = NA, **p-value = 0.04848**

| **Phenotype vs. Variant Type Residuals** | | | | |
|---|---|---|---|---|
| | Frameshift | Missense | Splice | Deletion |
| RCD | **2.67** | **-2.20** | -1.48 | 1.70 |
| CRD | -1.03 | -0.22 | 1.53 | -0.75 |
| RD | -0.64 | 0.27 | 0.66 | -0.75 |
| MD | **-2.49** | **3.41** | -0.06 | -0.57 |
| LCA | 1.68 | -0.85 | -1.09 | -0.20 |

**Cramér's V** = 0.1585 *Moderate

**Table S6. Phenotype vs. protein domain chi-square analysis.** Increased observed counts are highlighted in red, and decrease counts are shown in blue.

| **Phenotype vs. Protein Domain Observed** | | | | | | | | | | | |
|---|---|---|---|---|---|---|---|---|---|---|---|
| | N-Term | TM1 | CL1 | TM2 | EL1 | TM3 | CL2 | TM4 | EL2 | TM5 | C-Term |
| RCD | 10 | 0 | 2 | 3 | 15 | 3 | 2 | 2 | 25 | 1 | 2 |
| CRD | 2 | 1 | 0 | 0 | 6 | 3 | 4 | 0 | 15 | 2 | 1 |
| RD | 3 | 2 | 3 | 0 | 8 | 1 | 2 | 0 | 14 | 0 | 1 |
| MD | 4 | 0 | 0 | 0 | 5 | 0 | 1 | 0 | 11 | 0 | 1 |
| LCA | 0 | 0 | 0 | 0 | 0 | 0 | 0 | 0 | 3 | 0 | 0 |

| **Phenotype vs. Protein Domain Expected** | | | | | | | | | | | |
|---|---|---|---|---|---|---|---|---|---|---|---|
| | N-Term | TM1 | CL1 | TM2 | EL1 | TM3 | CL2 | TM4 | EL2 | TM5 | C-Term |
| RCD | 8 | 1 | 2 | 1 | 14 | 3 | 4 | 1 | 28 | 1 | 2 |
| CRD | 4 | 1 | 1 | 1 | 7 | 2 | 2 | 0 | 15 | 1 | 1 |
| RD | 4 | 1 | 1 | 1 | 7 | 2 | 2 | 0 | 15 | 1 | 1 |
| MD | 3 | 0 | 1 | 0 | 5 | 1 | 1 | 0 | 9 | 0 | 1 |
| LCA | 0 | 0 | 0 | 0 | 1 | 0 | 0 | 0 | 1 | 0 | 0 |

Monte Carlo Chi-Square
x-squared = 34.341 df = NA, p-value = 0.6767

| **Phenotype vs. Protein Domain Residuals** | | | | | | | | | | | |
|---|---|---|---|---|---|---|---|---|---|---|---|
| | N-Term | TM1 | CL1 | TM2 | EL1 | TM3 | CL2 | TM4 | EL2 | TM5 | C-Term |
| RCD | 1.09 | -1.46 | -0.05 | **2.09** | 0.40 | 0.09 | -1.19 | 1.70 | -0.97 | -0.28 | -0.05 |
| CRD | -1.24 | 0.50 | -1.19 | -0.92 | -0.62 | 1.41 | 1.72 | -0.75 | 0.14 | 1.92 | -0.08 |
| RD | -0.65 | 1.92 | **2.13** | -0.92 | 0.32 | -0.48 | 0.05 | -0.75 | -0.25 | -0.92 | -0.08 |
| MD | 0.96 | -0.70 | -0.91 | -0.70 | 0.15 | -1.09 | -0.25 | -0.57 | 0.71 | -0.70 | 0.40 |
| LCA | -0.65 | -0.24 | -0.32 | -0.24 | -0.92 | -0.38 | -0.43 | -0.20 | **2.01** | -0.24 | -0.32 |

**Cramér's V** = 0.0 *Negligible

**Table S7. Variant type vs. protein domain chi-square analysis.** Increased observed counts are highlighted in red, and decrease counts are shown in blue.

| **Variant Type vs. Protein Domain Observed** | | | | | | | | | | | |
|---|---|---|---|---|---|---|---|---|---|---|---|
| | N-Term | TM1 | CL1 | TM2 | EL1 | TM3 | CL2 | TM4 | EL2 | TM5 | C-Term |
| Frameshift | 10 | 0 | 5 | 1 | 22 | 6 | 5 | 2 | 48 | 3 | 1 |
| Missense | 4 | 3 | 1 | 0 | 8 | 0 | 3 | 0 | 9 | 1 | 2 |
| Splice | 10 | 0 | 0 | 2 | 11 | 2 | 2 | 0 | 26 | 0 | 2 |
| Deletion | 0 | 0 | 0 | 0 | 1 | 0 | 0 | 0 | 1 | 0 | 0 |

| **Variant Type vs. Protein Domain Expected** | | | | | | | | | | | |
|---|---|---|---|---|---|---|---|---|---|---|---|
| | N-Term | TM1 | CL1 | TM2 | EL1 | TM3 | CL2 | TM4 | EL2 | TM5 | C-Term |
| Frameshift | 13 | 2 | 3 | 2 | 23 | 4 | 5 | 1 | 45 | 2 | 3 |
| Missense | 4 | 0 | 1 | 0 | 7 | 1 | 2 | 0 | 14 | 1 | 1 |
| Splice | 7 | 1 | 2 | 1 | 12 | 2 | 3 | 1 | 24 | 1 | 1 |
| Deletion | 0 | 0 | 0 | 0 | 0 | 0 | 0 | 0 | 1 | 0 | 0 |

Monte Carlo Chi-Square

x-squared = 35.865, df = NA, p-value = 0.2574

| **Variant Type vs. Protein Domain Residuals** | | | | | | | | | | | |
|---|---|---|---|---|---|---|---|---|---|---|---|
| | N-Term | TM1 | CL1 | TM2 | EL1 | TM3 | CL2 | TM4 | EL2 | TM5 | C-Term |
| Frameshift | -1.29 | -1.89 | 1.47 | -0.72 | -0.23 | 1.22 | -0.26 | 1.31 | 0.79 | 0.85 | -1.54 |
| Missense | 0.06 | **3.97** | 0.03 | -0.77 | 0.56 | -1.27 | 1.21 | -0.63 | -1.83 | 0.48 | 1.46 |
| Splice | 1.49 | -1.11 | -1.58 | 1.46 | -0.42 | -0.24 | -0.63 | -0.90 | 0.58 | -1.29 | 0.56 |
| Deletion | -0.54 | -0.18 | -0.26 | -0.18 | 0.96 | -0.30 | -0.33 | -0.15 | 0.17 | -0.21 | -0.23 |

**Cramér's V** = 0.1006 *Weak
